# Appropriateness of the current parasitological control target for hookworm morbidity: a statistical analysis of individual-level data

**DOI:** 10.1101/2022.03.07.22271597

**Authors:** Veronica Malizia, Federica Giardina, Sake J. de Vlas, Luc E. Coffeng

## Abstract

**Background:** Soil-transmitted helminths affect almost 2 billion people globally. Hookworm species contribute to most of the related morbidity. Hookworms mainly cause anaemia, due to blood loss at the site of the attachment of the adult worms to the human intestinal mucosa. The World Health Organization (WHO) aims to eliminate hookworm morbidity by 2030 through achieving a prevalence of moderate and heavy intensity (M&HI) infections below 2%. In this paper, we aim to assess the suitability of this threshold to reflect hookworm-attributable morbidity.

**Methodology/Principal Findings:** We developed a hierarchical statistical model to simulate individual haemoglobin concentrations in association with hookworm burdens, accounting for low haemoglobin values attributable to other causes. The model was fitted to individual-level data within a Bayesian framework. Then, we generated different endemicity settings corresponding to infection prevalence ranging from 10% to 90% (0% to 55% M&HI prevalence), using 1, 2 or 4 Kato-Katz slides. For each scenario, we estimated the prevalence of anaemia due to hookworm. Our results showed that on average, haemoglobin falls below the WHO threshold for anaemia when intensities are above 2000 eggs per gram of faeces. For the different simulated scenarios, the estimated prevalence of anaemia attributable to hookworm ranges from 0% to 30% (95%-PI: 24% - 36%) being mainly associated to the prevalence of M&HI infections. Simulations show that a 2% prevalence of M&HI infections in adults corresponds to a prevalence of hookworm-attributable anaemia lower than 1%.

**Conclusions/Significance:** Our results support the use of the current WHO thresholds of 2% prevalence of M&HI as a proxy for hookworm morbidity. A single Kato-Katz slide may be sufficient to assess the achievement of the morbidity target. Further studies are needed to elucidate haemoglobin dynamics pre- and post-control, ideally using longitudinal data in adults and children.

**Author summary:** Soil-transmitted helminths affect almost 2 billion people globally. About 65% of the related morbidity is attributable to hookworm infection, especially anaemia, which is caused by adult worms attaching to the intestinal mucosa of the human host to feed on blood. The World Health Organization (WHO) defines the target for the elimination of morbidity as reaching moderate-to-heavy intensity infections prevalence < 2%. In this paper, we aim to assess the suitability of the current WHO threshold to reflect hookworm-attributable anaemia. To this end, we developed a statistical model to simulate individual haemoglobin concentrations in association with hookworm burdens in adults, accounting for low haemoglobin values attributable to other causes, such as malnutrition and malaria. We used model outcomes to estimate the prevalence of hookworm-attributable anaemia in different endemicity scenarios. Our predictions suggested that anaemia is mainly associated to moderate-to-heavy intensity infection and that the 2% threshold corresponds to a prevalence of hookworm-attributable anaemia <1%. These results support the use of the current WHO threshold as a proxy for hookworm morbidity. Our predictions represent a first step towards better quantifying the prevalence of anaemia due to hookworm infection.

## Introduction

Soil-transmitted helminths (STH) are estimated to affect almost 2 billion people globally [1], with the vast majority living in developing countries. Transmitted through contaminated soil, STH are responsible for considerable morbidity. About 65% of the overall estimated STH morbidity is attributable to hookworm infection, which is commonly caused by the helminth parasites *Necator americanus* and *Ancylostoma duodenale* [2]. The most common condition associated with hookworm infection is anaemia resulting from adult hookworms attaching to the mucosa of the small intestine in the human host to feed on blood. The uptake of blood and the extra loss due to wounds left by detaching worms reduce haemoglobin (Hb) and iron levels of the host, until the nutritional iron reserves and intake are not sufficient anymore [3]. Hookworm infection has been shown to significantly contribute to the global burden of anaemia in many countries [4].

The World Health Organization (WHO) categorises hookworm infections as light (≤ 1999 eggs per gram of faeces (epg)), moderate (2000 – 3999 epg) and heavy (≥ 4000 epg) [5]. The current global target set by the WHO is the elimination of hookworm as a public health problem by 2030, defined as reaching a moderate-to-heavy intensity (M&HI) prevalence < 2% in school-aged children [6]. Although anaemia is thought to be a consequence of M&HI infection, lower Hb levels in adults have been associated with all levels of infection intensity [7]. The number of adult worms in the human host that are sufficient to cause anaemia depends on several factors, such as the hookworm species causing the infection, the underlying nutritional reserves of the host, and coinfection with other parasites [8]. Clear evidence for the definition of the WHO guidelines and the use of M&HI prevalence of infection as indicator of hookworm morbidity is currently lacking.

Previous studies estimated the effect of hookworm infection intensity on blood iron reserves [3] and investigated the relationship between Hb and faecal egg counts [9]. For instance, Lwambo et al. [9] showed that Hb significantly declines for infection intensities above 2000 epg. They also predicted a non-linear relationship between prevalence of infection and prevalence of hookworm-related anaemia where the latter steeply increases with high prevalence of infection. In these studies, results are presented in terms of overall prevalence of anaemia in the population. However, in most of the settings where hookworm infection is prevalent, malnutrition and coinfection with other endemic infectious diseases (e.g., malaria, schistosomiasis, other neglected tropical diseases) are also common. Therefore, it is important to account for an underlying prevalence of anaemia in the population due to causes other than hookworm infection.

The aim of this paper is to assess the suitability of the current WHO targets using prevalence of M&HI infection as a proxy for hookworm morbidity. To do this, we estimate the prevalence of anaemia attributable to hookworm infection as a function of hookworm prevalence (M&HI and any intensity). We do so by modelling how individual level Hb concentrations change in response to individual hookworm burden, indirectly observed through excreted egg counts. We account for anaemia attributable to other causes by assuming an Hb distribution that allows low values also in absence of worms. To extend this approach to other settings, we generate different endemicity scenarios and for each we estimate the prevalence of anaemia attributable to hookworm. We further consider the impact of different sampling schemes for diagnosis using the Kato-Katz (KK) thick smear technique on the evaluation of the morbidity target.

## Methods

### Data

We analysed individual data from a rural community in Uganda (N = 2037 individuals), consisting of Hb values and egg count measurements for hookworm and *Schistosoma mansoni* detection, collected by means of two KK slides per day per individual, for a total of two days and four slides for each disease. Age, sex, and geographical location were also recorded. These data have been previously analysed to estimate the spatial and genetic variation in intensity of hookworm infection [10]. The original study protocol was approved by the Makerere University Faculty of Medicine Research and Ethics Committee (#2008-043), the Uganda National Council of Science and Technology (#HS 476) and the London School of Hygiene and Tropical Medicine Ethics Committee (#5261) [10]. Hb concentrations and egg counts for hookworm were available for 1840 individuals. The WHO provides the definition of anaemia based on age- and sex-specific thresholds [11]: women and men > 15 years old are defined anaemic if their Hb concentration is below 120 and 130 g/L, respectively. Due to high age/sex-variation in anaemia thresholds for children and the assumption that exposure to hookworm infection stabilises in adult age [12], we focused our analysis on the adult population (≥ 20 years, both sexes). In ten individuals, extremely high egg counts were recorded (up to a maximum of 35,000 epg). We hypothesised that these few individuals were infected with the hookworm species *A. duodenale* and therefore excluded them (individuals with a mean egg count > 10,000 epg) from the analysis. The final population is composed of N=695 adults with a 52% prevalence of any intensity of hookworm infection, 4% prevalence of M&HI of infection and 41% prevalence of overall anaemia. The process of data selection is summarised in **Fig 1**.

**Fig 1.**
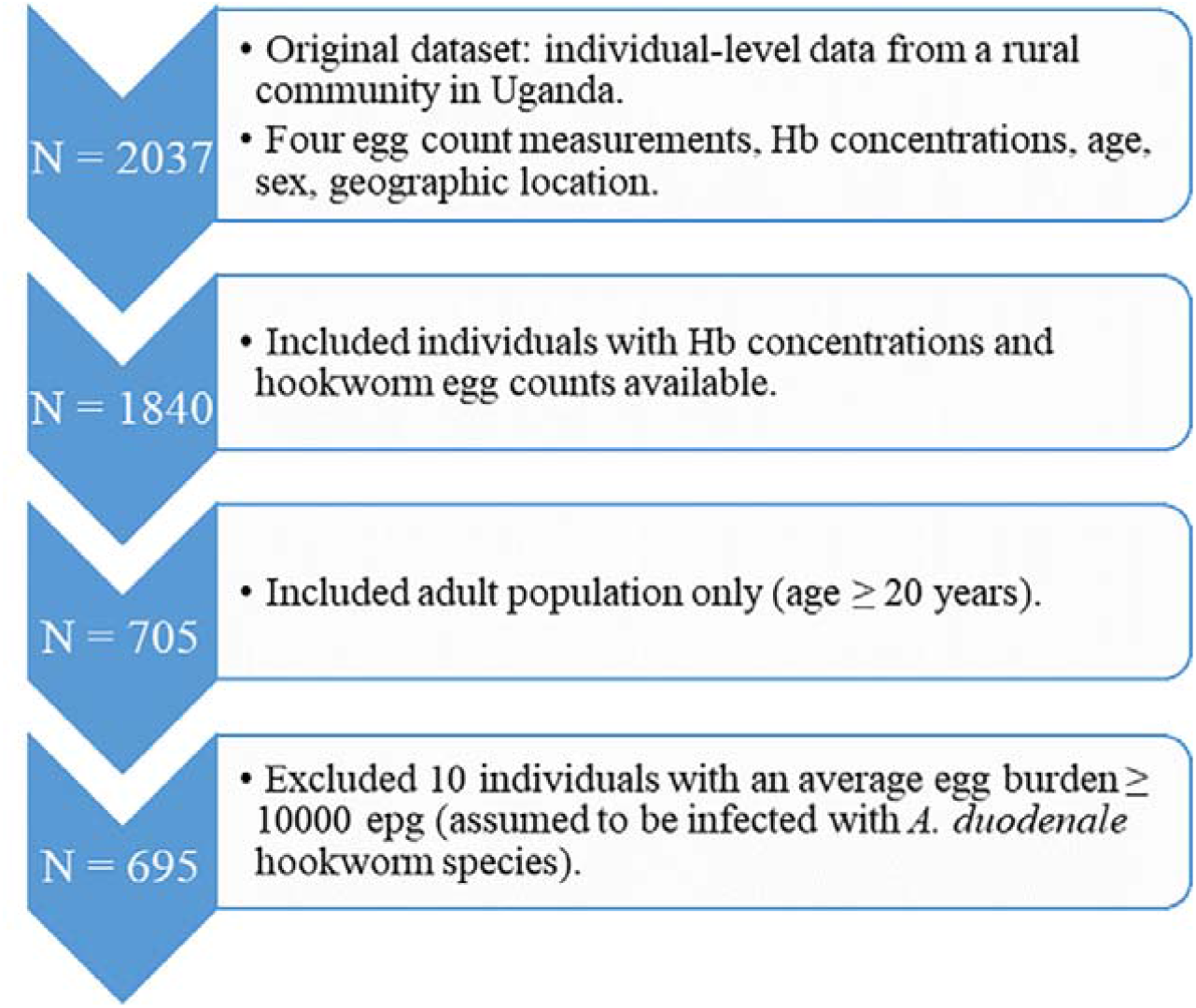
Flowchart describing the process of data selection for the statistical analysis.

### Model

The association between Hb values and egg counts was studied by means of a hierarchical statistical model defined at individual level, with a latent variable representing the unobserved worm burdens in each human host. The distribution of worm burdens in a population is known to be highly over-dispersed, with most people harbouring no or a small number of worms and only few highly infected individuals. The parasitological part of the hierarchical model is described as follows:

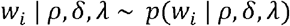

where *w*_*i*_ is a semi-continuous latent variable representing the worm burden for the *i*-th individual. Its probability density function is defined as:

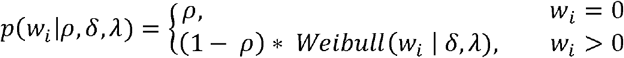

The latent variable *w*_*i*_ was thus modelled from a finite mixture of a Bernoulli distribution with probability ρ (i.e., the probability of having zero worms) and a Weibull distribution with shape parameter δ and scale λ. The employment of this semi-continuous variable is meant to approximate the discrete negative binomial distribution, which has commonly been employed for describing over-dispersed worm burdens in human populations. This approximation was necessary to be able to use the Hamiltonian Monte Carlo algorithm to sample from the posterior distribution, which does not allow the use of a discrete latent variable. The latent variable *w*_*i*_ enabled us to discriminate between infected (if *w*_*i*_ > 0) and uninfected individuals (if *w*_*i*_ = 0).

The corresponding individual egg counts are simulated in the model as follows:

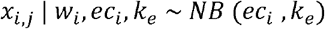

The variable *x*_*i,j*_ is the *j*^*th*^ egg count for the *i*-th individual, with *j* = 1, …, 4. The parameters *ec*_*i*_ and *k*_*e*_ represent the expectation and the aggregation parameter of the egg count distribution. The aggregation parameter *k*_*e*_ encapsulates the variability within the same stool specimen and between different stool samples. The expected individual egg burden was assumed to depend on the underlying worm load *w*_*i*_ according to a hyperbolic saturating function [12] describing density dependent fecundity. The function is defined as follows: 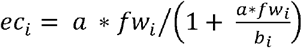 where *fw*_*i*_ = *w*_*i*_/2 approximates the female worm load, the parameter *a* is the average egg production for each female worm in absence of density dependence effects, and *b*_*i*_ the maximum host egg output. The latter was assumed to vary between hosts due to host suitability for infection and was assumed to follow a gamma distribution with mean β representing the population average saturation level in egg production. The values of *a* and the shape of the gamma distribution (which dictates the level of variation in host suitability) were set to values available from previous work (**Table 1**).

**Table 1.** Parameters employed in the model. The parameters can be either given in input or estimated from the model. For input parameters the corresponding value is reported, the prior distribution for the Bayesian estimation otherwise. When available, the source of the information is cited. Prior distributions are parameterised in terms of mean and standard deviation (normal) or scale (Cauchy).

| Parameter | Origin | Value | Prior | Source |
| --- | --- | --- | --- | --- |
| Probability of having zero worms ($\rho \in [0, 1]$) | Estimated | - | $\rho \sim \text{Uniform}(0, 1)$ | - |
| Shape of the Weibull distribution for worm burdens in infected individuals ($\delta > 0$) | Estimated | - | $\delta \sim \text{Normal}^+(0.5, 1)$ | - |
| Scale of the Weibull distribution for worm burdens in infected individuals ($\lambda > 0$) | Estimated | - | $\lambda \sim \text{Cauchy}^+(2, 1)$ | - |
| Initial slope of the egg production function ($a$) | Input | 200 epg | - | [12] |
| Mean saturation level of the egg production function ($\beta$) | Input | 10,000 epg | - | Pre-set |
| Shape and rate of the gamma distribution for individual relative susceptibility to infection | Input | 50 | - | [12] |
| Aggregation parameter for egg counts ($k_e > 0$) | Estimated | - | $k_e \sim \text{Normal}^+(0, 2)$ | [12] |
| Logarithm of geometric mean of Hb [g/L] distribution in uninfected individuals ($\mu_0$) | Estimated | - | $\mu_0 \sim \text{Normal}(\log(129), 0.1)$ | [13, 14] |
| Scale parameter of Hb distribution ($\sigma > 0$) | Estimated | - | $\sigma \sim \text{Normal}^+(0, 0.5)$ | [13, 14] |
| Steepness of the logistic decrease ($\varphi_1 > 0$) | Estimated | - | $\varphi_1 \sim \text{Normal}^+(1, 3)$ | - |
| Location of the sigmoid<br>($\varphi_0 > 0$) | Estimated | - | $\varphi_0 \sim \text{Normal}^+(6, 2)$ | - |

<sup>+</sup> The prior distribution is left-truncated at zero.

The mean saturation level β was set to 10,000 epg to assure the model was able to catch the entire range of observed egg counts.

We further assumed that Hb concentrations (*y*_*i*_) follow a LogNormal distribution with standard deviation σ representing the natural variation between individuals and a mean that depends on an individual’s infection status. In absence of hookworm infection (i.e. *w*_*i*_ = 0), we assumed a mean of *µ*_0_. For individuals with a worm burden w_*i*_ > 0, we assumed the expected Hb to follow a logistic decrease on the logarithmic scale with increasing worm burden, i.e.:

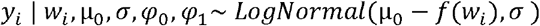

where the logistic decrease is defined as 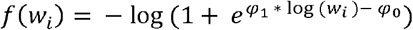. Here, *φ*_1_ represents the steepness of the decreasing function and *φ*_0_ is proportional to the *x-*axis intercept of the middle point of the sigmoid. The full list of employed parameters is presented in **Table 1**.

The model was fitted to the data within a Bayesian framework. The probability ρ of having zero worms was assigned a Uniform prior between 0 and 1. The hyper-parameters of the Weibull distribution modelling the individual worm burdens in infected individuals were assigned vague hyper-priors. Prior distributions for parameters *µ*_0_. and σ were based on information available from the literature [13, 14]. Weakly informative priors were assigned to *φ*_0_, *φ*_1_ and *k*_e_. Prior information on *φ*_1_ and *k*_e_ is limited to mathematical constraints.

Posterior distributions of the estimated parameters and predictions were simulated in Stan [15] (code for model specification in Stan is publicly available [16]), a probabilistic programming language for statistical modelling. Stan allows the user to perform full Bayesian statistical inference using Markov Chain Monte Carlo (MCMC) sampling (based on NUTS or HMC sampling algorithm). Simulations were run and further analysed in R (version 3.6.1) [17], using the package *rstan* [18]. The algorithm was run on 4 independent Markov chains, each with 10,000 draws, for a total of 20,000 post-warmup draws (the initial 5,000 draws per chain were considered for warm-up). Convergence was assessed by visual examination of the trace plots and the convergence diagnostic methods provided by Stan. Fit evaluation was performed by means of full posterior predictive checks [19]. In this procedure, the population was replicated for each draw of the joint posterior distribution of model parameters and then compared to the original data.

### Simulations

After fitting the model, the posterior distributions obtained for each of the fitted parameters were used to study the appropriateness of the current WHO parasitological target as proxy for hookworm morbidity. We simulated different endemicity scenarios characterised by a specific worm distribution in the population (i.e., a specific combination of the three parameters ρ, δ and λ describing the mixture distribution of worm burdens) resulting in different prevalence of hookworm infection (any intensity and M&HI) settings. The assumptions about the distributions that describe the actual egg counts, the Hb concentrations and the background risk of anaemia in the simulated populations were the same as assumed while fitting the observed data. The simulated infection prevalence ranged from 10% to 90% (any intensity) and 0% to 55% (M&HI). For each scenario, we generated a population of 1000 individuals using a random sample of 5000 draws from the posterior distribution of the parameters and we computed: i) the KK-based hookworm prevalence of any infection, ii) the KK-based prevalence of M&HI infection, iii) overall prevalence of anaemia, iv) anaemia attributable to hookworm infection. The latter is defined as the difference between the overall predicted anaemia and a baseline anaemia in the counterfactual scenario without worm infections, estimated from the model. This computation does not explore the contribution of hookworm infection to the severity of anaemia. Each of the aforementioned quantities were computed based on 3 different sampling schemes to assess the KK-based prevalence of hookworm: i) a single KK slide, which is the most common diagnostic scheme in the field, ii) duplicate slides, and iii) four repeated slides, in line with the dataset used in this study.

All analyses were performed in accordance with the Policy-Relevant Items for Reporting Models in Epidemiology of Neglected Tropical Diseases (PRIME-NTD) criteria.[20] (**S1 Table**). The full model specification and the code for data selection, statistical analysis and simulations are publicly available.[16]

## Results

### Association between individual haemoglobin levels and egg counts

Our statistical model fitted the individual data from Uganda well, which suggests that a logistic function is a reasonable representation of the effect of increasing hookworm burden on Hb levels in adults (**Fig 2**).

**Fig 2.**
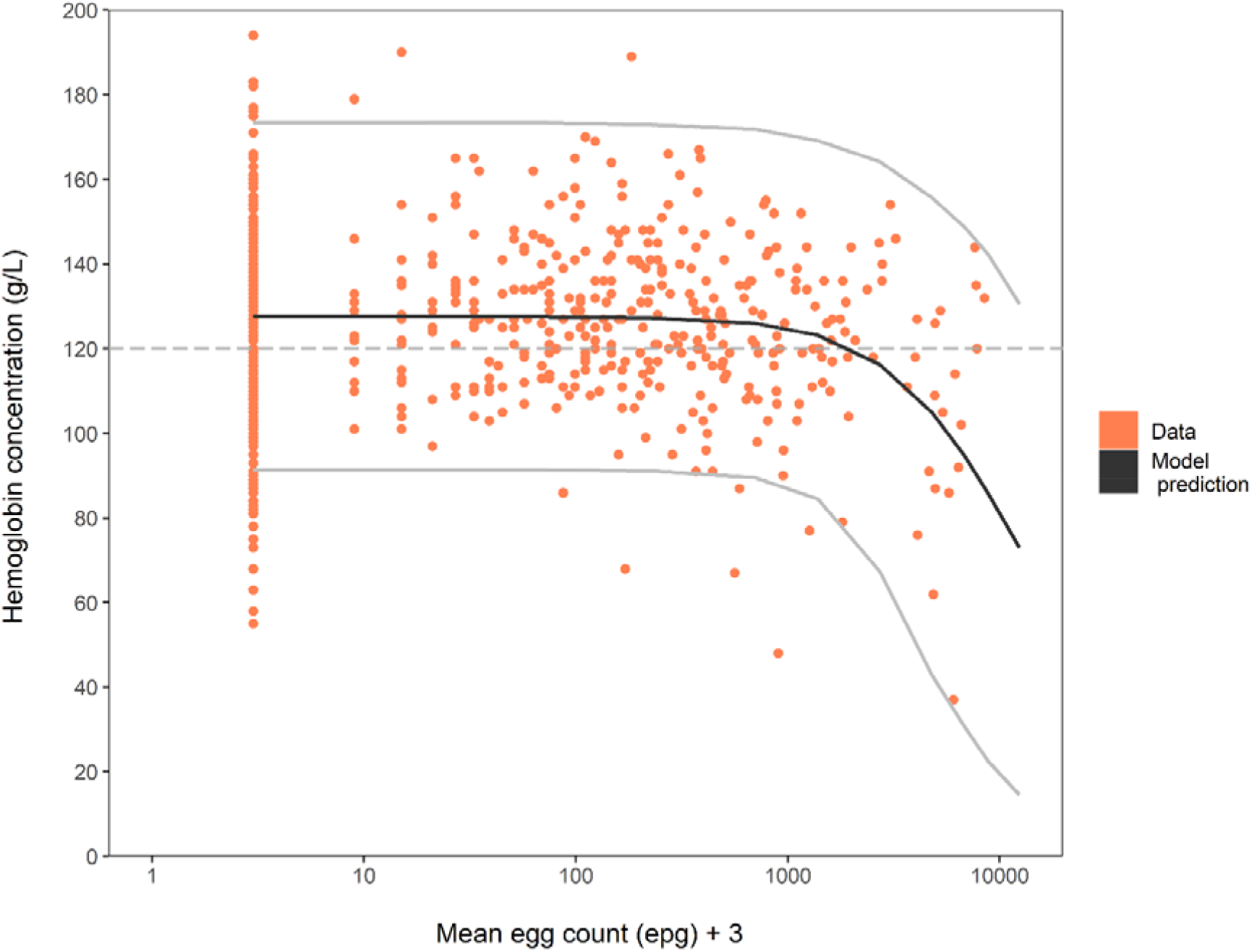
Relationship between individual haemoglobin concentrations (g/L) and mean egg count (epg). Orange bullets represent observed individual data used for model fit. The joint posterior predictive distribution of Hb and mean egg count based on 20,000 replications is displayed through the mean of the Hb predictions (y-axis) and the geometric mean of egg count predictions (x-axis) (black line). Mean egg count data are shifted of +3 epg (i.e. half of the detection limit) to display the zero counts on the logarithmic-scaled axes. The 95% Bayesian Credible Interval (light grey lines) is shown for the marginal Hb posterior predictive distribution. The WHO threshold defining anaemia used in the model is displayed as horizontal dashed line.

This figure shows the joint posterior predictive distribution of Hb and egg counts based on 20,000 iterations, displayed via the mean of Hb predictions (y-axis) and the geometric mean of egg count predictions (x-axis). The 95% Bayesian Credible Interval (BCI) is solely displayed for the marginal Hb posterior predictive distribution. In absence of hookworm infection, Hb levels in the population are distributed with an estimated geometric mean of 126 g/L (95%-BCI: 124.3 g/L – 127.5 g/L) and a standard deviation of the Hb natural logarithm equal to 0.16 (95%-BCI: 0.15 – 0.17). The estimated parameters of the logistic function showed that hookworm infection has little effect on Hb levels in the population for intensities up to 1000 epg and that Hb falls below the WHO threshold for anaemia when intensities are above 2000 epg. See **Table 2** for detailed parameter estimates.

**Table 2.** Model parameter estimates. Estimates represent posterior distributions of model parameters. For each parameter, Rhat can be used as indicator for Markov chains convergence (at convergence, Rhat=1). BCI = Bayesian credible interval, based on the central 95% percentiles of the posterior draws.

| Parameter | Mean [95%-BCI] | Rhat |
| --- | --- | --- |
| Probability of having zero worms ($\rho \in [0, 1]$) | 0.38 [0.32, 0.43] | 1.001 |
| Shape of the Weibull distribution for worm burdens in infected individuals ($\delta > 0$) | 0.48 [0.40, 0.55] | 1.001 |
| Scale of the Weibull distribution for worm burdens in infected individuals ($\lambda > 0$) | 3.31 [2.34, 4.41] | 1.001 |
| Aggregation parameter for egg counts ($k_e > 0$) | 0.66 [0.59, 0.74] | 1.000 |
| Logarithm of geometric mean of Hb [g/L] distribution in uninfected individuals ($\mu_0$) | 4.84 [4.82, 4.85] | 1.000 |
| Scale parameter of Hb distribution ($\sigma > 0$) | 0.16 [0.15, 0.17] | 1.000 |
| Steepness of the logistic decrease ($\varphi_1 > 0$) | 1.90 [1.33, 2.54] | 1.001 |
| Location of the sigmoid ($\varphi_0 > 0$) | 9.23 [6.96, 11.90] | 1.000 |

The posterior predictive checks obtained from the model fit to the data recorded a prevalence of hookworm infection in the population of 54% (95%-BCI: 48%-59%; 52% in data), a M&HI infection prevalence of 4.5% (95%-BCI: 3% - 7%; 4% in data) and a prevalence of overall anaemia of 40% (95%-BCI: 36% - 45%; 41% in data). The posterior distributions of the parameters *μ*_0_ and σ suggested a baseline prevalence of anaemia (in absence of hookworm infection) in the population of 38% (95%-BCI: 35% - 41%). This represents the prevalence of anaemia that may be attributed to other causes.

Full posterior predictive checks for Hb levels and egg counts distributions, based on 20,000 replicated datasets were compared to the original data and used to assess model fit (**Fig 3**). The posterior predictive distributions approximated the observed egg counts and Hb distributions in the population reasonably well.

**Fig 3.**
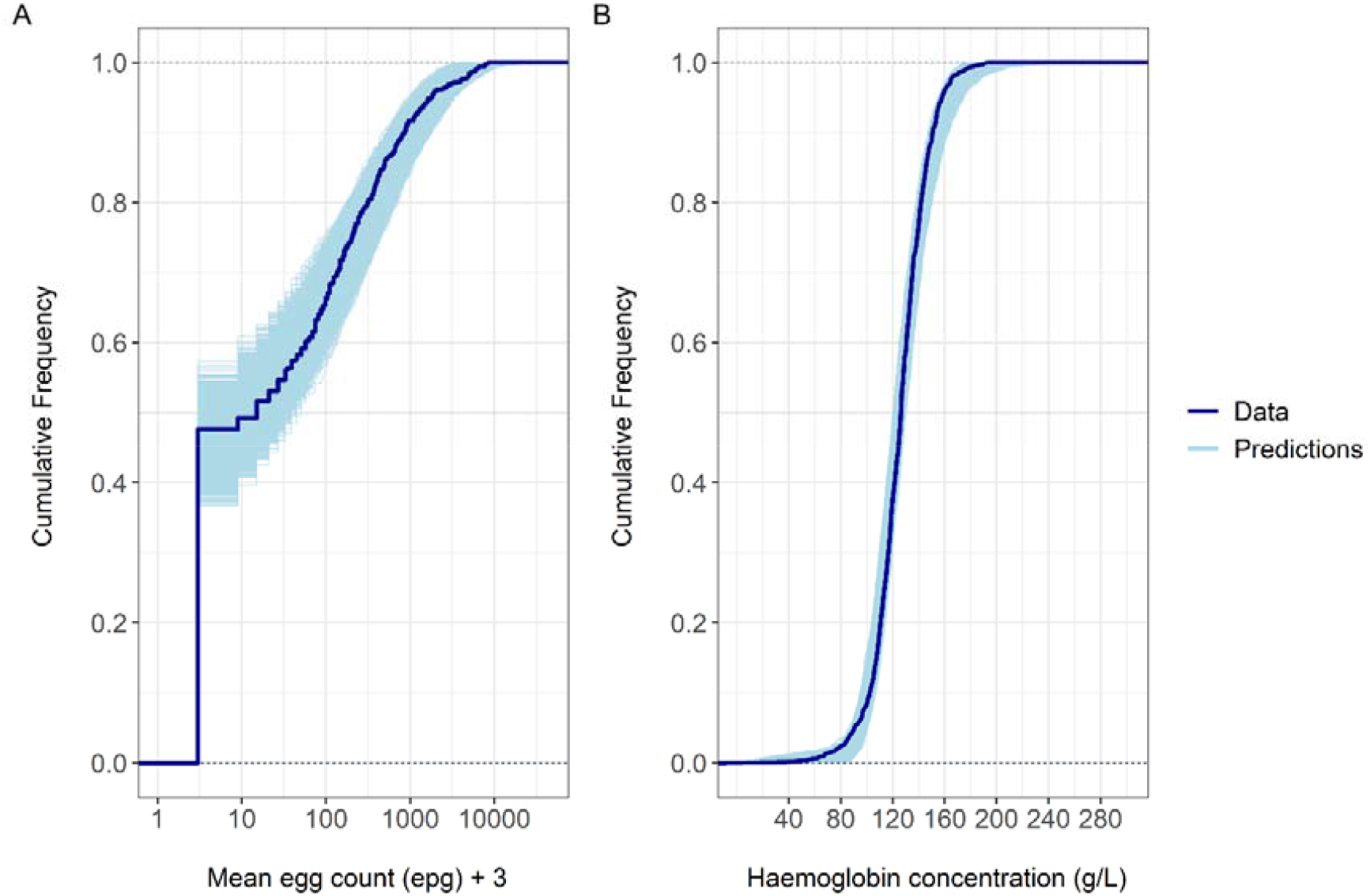
Comparison of observed data and model prediction. (A) Full posterior predictive checks for the mean egg counts. Mean egg count data are shifted of +3 epg (i.e. half of the detection limit) to display the zero counts on the logarithmic-scaled axes. (B) Full posterior predictive checks for haemoglobin concentrations. Dark lines represent the corresponding distribution observed in the Ugandan population. Both marginal predictive distributions are displayed through the cumulative distribution function of the predictions based on 20,000 replications (light lines) and compared with observed data.

### Simulated prevalence of anaemia due to hookworm infection

The posterior distributions obtained for each of the fitted parameters were used for simulating different endemicity scenarios, assuming that the risk of anaemia due to causes other than hookworm infection is the same as in the Ugandan data. The simulated prevalence of anaemia attributable to hookworm is displayed in **Fig 4** with its mean and corresponding 95% prediction interval (PI), as a function of infection prevalence (any intensity and M&HI). The figure shows that the prevalence of anaemia attributable to hookworm ranged from 0% to 25% (95%-PI: 20% - 30%) by varying endemicity scenarios and it showed strong association with the prevalence of M&HI infections. In all the simulated scenarios, a 2% prevalence of M&HI infections in adults corresponded to a prevalence of anaemia attributable to hookworm infections lower than 1%.

**Fig 4.**
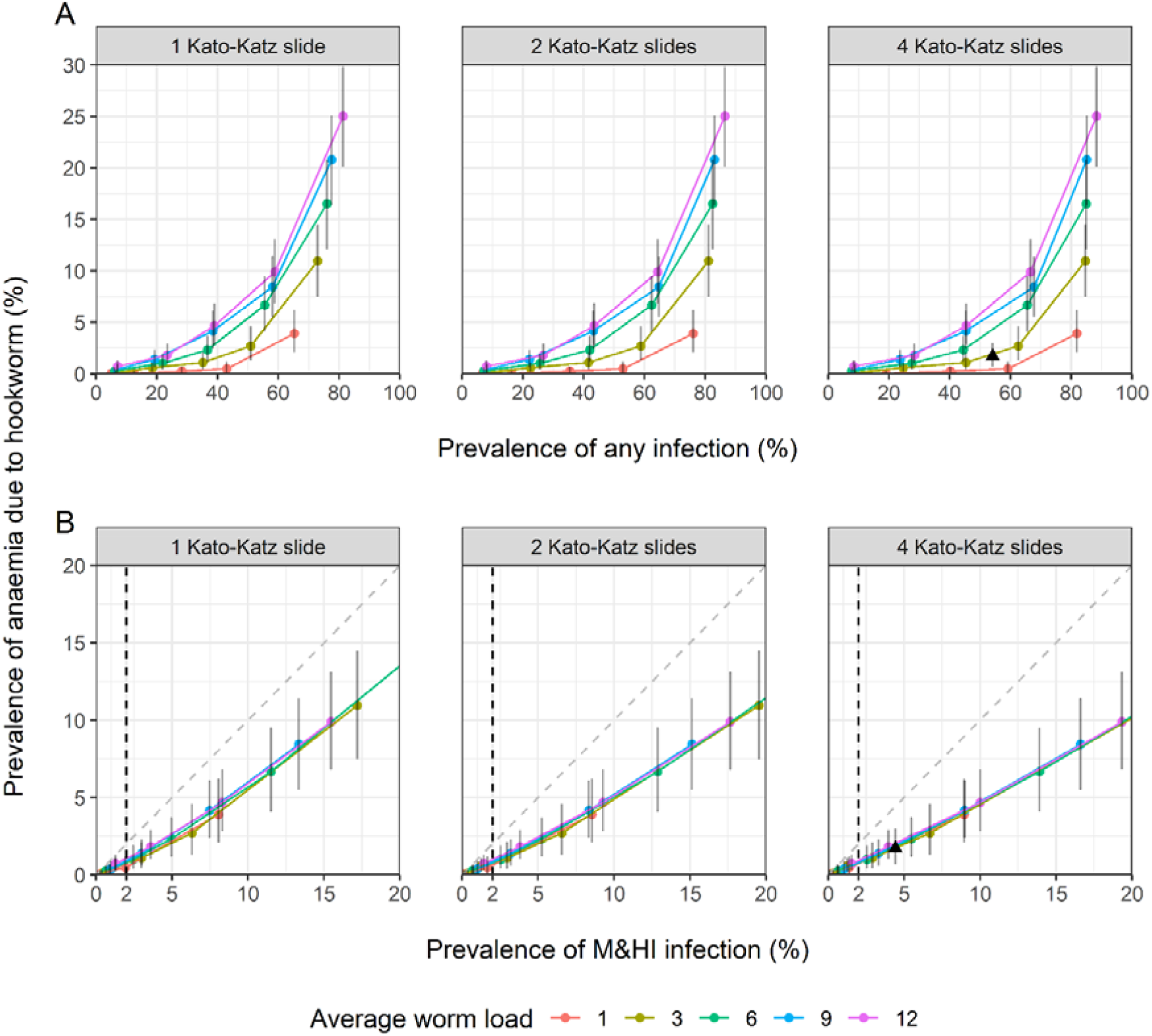
Estimated prevalence of anaemia attributable to hookworm in adult populations, for different infection prevalence scenarios. Predictions are displayed for different infection prevalence scenarios, defined by varying the probability ρ of having zero worms among the five values 0.1, 0.3, 0.5, 0.7, 0.9 (different bullets of the same colour) and the population mean worm burden (different colours). The shape parameter of the worm distribution across infected individuals is fixed to δ = 0.5 (as estimated from the Ugandan data). From left to right, the prevalence of any hookworm infection (A) and moderate-to-heavy intensity of infection (B) is based on 1, 2, 4 Kato-Katz slides, respectively. Each point (x, y) represents the mean predictions of infection prevalence (x-value) vs. hookworm-anaemia prevalence (y-value). The error band reports the 95% prediction interval (PI) of predicted hookworm-anaemia. The black triangle indicates the predictions (mean and 95%-PI) for the scenario resulting from the fit to the Ugandan dataset. Dashed vertical lines mark the WHO morbidity threshold of 2% moderate-to-heavy intensity prevalence. The bisector in panels (B) is added to visualise the robustness of the prevalence of moderate-to-heavy intensity of infection over the three sampling schemes.

Our simulations showed that the prevalence of infection is sensitive to the number of KK simulated measurements. However, low values of M&HI infection prevalence (<15%) in a given scenario were relatively consistent over the three sampling schemes (1, 2 or 4 KK simulated measurements).

The endemicity scenarios presented in **Fig 4** are based on a fixed value of the shape parameter for the distribution of worms across infected individuals (*δ* = 0.5, i.e., approximately the value estimated for the Ugandan dataset). Simulations for alternative values of the shape parameter resulted in a similar pattern, with the prevalence of anaemia attributable to hookworm ranging from 0% to 30% (95%-PI: 24% - 36%), by varying endemicity scenarios (**S1 Fig**).

## Discussion

The aim of this study was to estimate the prevalence of anaemia attributable to hookworm infection and to evaluate whether the current guidelines for hookworm control properly reflect actual hookworm morbidity. To that end, we used a statistical hierarchical model that estimates the mechanisms of Hb decrease in response to increasing worm burden at individual level. Our analysis suggests that the estimated hookworm-attributable anaemia is strongly associated to M&HI infections. The current threshold used as indicator for elimination of hookworm morbidity (2% of M&HI prevalence as for the 2030 WHO target) corresponds to less than 1% prevalence of anaemia due to hookworm in the simulated endemicity settings. Therefore, our predictions confirm that the current threshold adopted by the WHO for the elimination of hookworm as a public health problem seems suitable. Even though our results are solely based on Hb values and egg counts from adults we expect similar predictions for the anaemia attributable to hookworm in children. Adults can be infected with hookworm to the same extent as children. Adults were shown to be responsible of most of the exposure to hookworm infection, with higher experienced morbidity [21].

Hookworm infections were found to have little effect on Hb levels for intensities up to 1000 epg. In fact, it is known that the uptake of blood by few worms should be easily counterbalanced by stored iron in the host, with some variation depending on individual nutritional reserves [3]. Our analysis show that for moderate (≥ 2000 epg) and heavy (≥ 4000 epg) intensity of infection the predicted average Hb drastically drops below the WHO threshold for anaemia. These results are in line with Lwambo *et al*. [9].

The simulated scenarios also show that the predicted KK based prevalence is dependent on the number of repeated measurements, but values of M&HI infection prevalence < 15% are quite robust over the three simulated sampling schemes (1, 2 or 4 KK) (**Fig 4**). Recent work by our team [22] reached similar conclusions, suggesting that 1 slide/sample may be sufficient for determining prevalence of M&HI infection to assess the morbidity target.

Previous modelling studies estimated the prevalence of hookworm morbidity from existing data [3, 9, 23] without explicitly considering anaemia due to other causes. In our model we accounted for an underlying prevalence of anaemia due to other causes such as nutritional factors or other infectious diseases, by explicitly modelling individual Hb variation. This approach finds support from a recently published systematic review and meta-analysis [24] which showed that the intensity of hookworm infection and the influence of other sources on the risk of anaemia drive differences in mean Hb concentrations, in hookworm endemic settings.

The Ugandan dataset used to fit our model showed some peculiarities we had to account for in our analysis. The subset of population detected with zero eggs for hookworm reported relatively low average Hb, presumably indicating other abundant sources of anaemia: part of the population was co-infected with *Schistosoma mansoni* [10]. The majority of individuals who tested positive to hookworm was lightly infected (only 4% of M&HI infection prevalence). Therefore, we did not include sex differences in the model, with the aim of gathering enough information about the steepness of the Hb decrease. However, we expect male and female population to show the same decreasing dynamics in response to increasing worm burdens, considering biological differences in how Hb concentrations are distributed in absence of infection between the two groups. The analysed Ugandan population is also heterogeneous with respect to the hookworm species causing the infection. In light of considerable differences in egg production [8], density-dependent fecundity effects [25] and in contribution to morbidity, we excluded ten individuals that we assumed to be infected with *A. duodenale* (detected epg ≥ 10,000). However, few individuals infected with *A. duodenale* at lower intensities could not be identified and could well still be part of the analysed data, contributing to a milder estimated slope of the Hb logistic decrease. We nevertheless do not expect those data to have a critical impact on the overall estimation of the relationship between Hb and worm burdens.

The distribution of worms between individuals is commonly described by a negative binomial distribution, parameterised with a mean worm burden and an aggregation parameter regulating the level of over-dispersion. However, we coded the model in Stan [15] which does not allow the use of a discrete latent variable [15]. Therefore, we used a semi-continuous approximation resulting from a finite mixture of a Weibull distribution and a point mass at zero, that can be interpreted as the level of true infection intensity in the host. The employment of a finite mixture distribution for modelling worm burdens allows discrimination between infected and uninfected individuals, providing a prediction of the true prevalence of infection (i.e., presence of worms). In our model we account for possible false negative results from the KK test, that we assume to be ascribed to an undetectable level of infection (low worm loads) or to the sensitivity of the test (simulated through the over-dispersion parameter of the negative binomial distribution used for modelling egg counts).

In conclusion, this study represents a first step towards better quantifying the prevalence of anaemia due to hookworm infection. Our results support the use of the current WHO threshold of 2% prevalence of M&HI as a proxy for hookworm morbidity. In addition, a single KK slide may be sufficient for assessing this morbidity target. Further studies are needed to elucidate Hb dynamics pre- and post-mass drug administration, where the infection harbouring time plays a crucial role [24], ideally using longitudinal data collected in both children and adults.

## Data Availability

All data and files containing full model specification, the code for data selection, statistical analysis and simulations are available from the "Hookworm anaemia model" public repository (https://github.com/VeronicaMalizia/Hookworm_anaemia_statistical_model.git).

## Authors’ contributions

**Conceptualization:** Veronica Malizia, Federica Giardina, Sake J. de Vlas, Luc E. Coffeng.

**Data curation:** Veronica Malizia, Luc E. Coffeng.

**Formal analysis:** Veronica Malizia, Federica Giardina, Luc E. Coffeng.

**Funding acquisition:** Sake J. de Vlas, Luc E. Coffeng.

**Methodology:** Veronica Malizia, Federica Giardina, Luc E. Coffeng.

**Software:** Veronica Malizia.

**Supervision:** Federica Giardina, Sake J. de Vlas, Luc E. Coffeng.

**Validation:** Veronica Malizia, Federica Giardina, Luc E. Coffeng.

**Visualization:** Veronica Malizia, Federica Giardina., Luc E. Coffeng.

**Writing – original draft:** Veronica Malizia, Federica Giardina.

**Writing – review & editing:** Veronica Malizia, Federica Giardina, Sake J. de Vlas, Luc E. Coffeng.

## Acknowledgments

The authors gratefully acknowledge Professor Simon J Brooker (Bill & Melinda Gates Foundation, Seattle, WA, USA), Dr. Rachel L Pullan (Department of Disease Control, London School of Hygiene and Tropical Medicine, London, United Kingdom) and Dr. Narcis B. Kabatereine (Vector Control Division, Ministry of Health, Kampala, Uganda) for collecting and sharing the data. The authors would like to thank Professor Bruno Levecke (Ghent University, Faculty of Veterinary Medicine, Merelbeke, Belgium) for the useful discussions about the differences in egg production and contribution to morbidity among hookworm species.

## Supporting information

**S1 Fig. Estimated prevalence of anaemia attributable to hookworm in adult populations, for different infection prevalence scenarios**. Predictions are displayed for different infection prevalence scenarios, defined by varying the probability ρ of having zero worms among the five values 0.1, 0.3, 0.5, 0.7, 0.9 (different points of the same colour), the population mean worm burden (different colours) and the shape parameter δ of worm distribution in infected individuals (different shapes). From left to right, the prevalence of any hookworm infection (A) and moderate-to-heavy intensity of infection (B) is based on 1, 2, 4 Kato-Katz slides, respectively. Each point (x, y) represents the mean predictions of infection prevalence (x-value) vs. hookworm-anaemia prevalence (y-value). The error band reports the 95% Confidence Interval (CI) of predicted hookworm-anaemia. Dashed vertical lines mark the WHO morbidity threshold of 2% moderate-to-heavy intensity prevalence. The bisector in panels (B) is added to visualise the robustness of the prevalence of moderate-to-heavy intensity of infection over the three sampling schemes.

**S1 Table. The Policy-Relevant Items for Reporting Models in Epidemiology of Neglected Tropical Diseases (PRIME-NTD)**.

