## Supplementary figures and images for "Appropriateness of the current parasitological control target for hookworm morbidity: a statistical analysis of individual-level data"

### S1 Fig

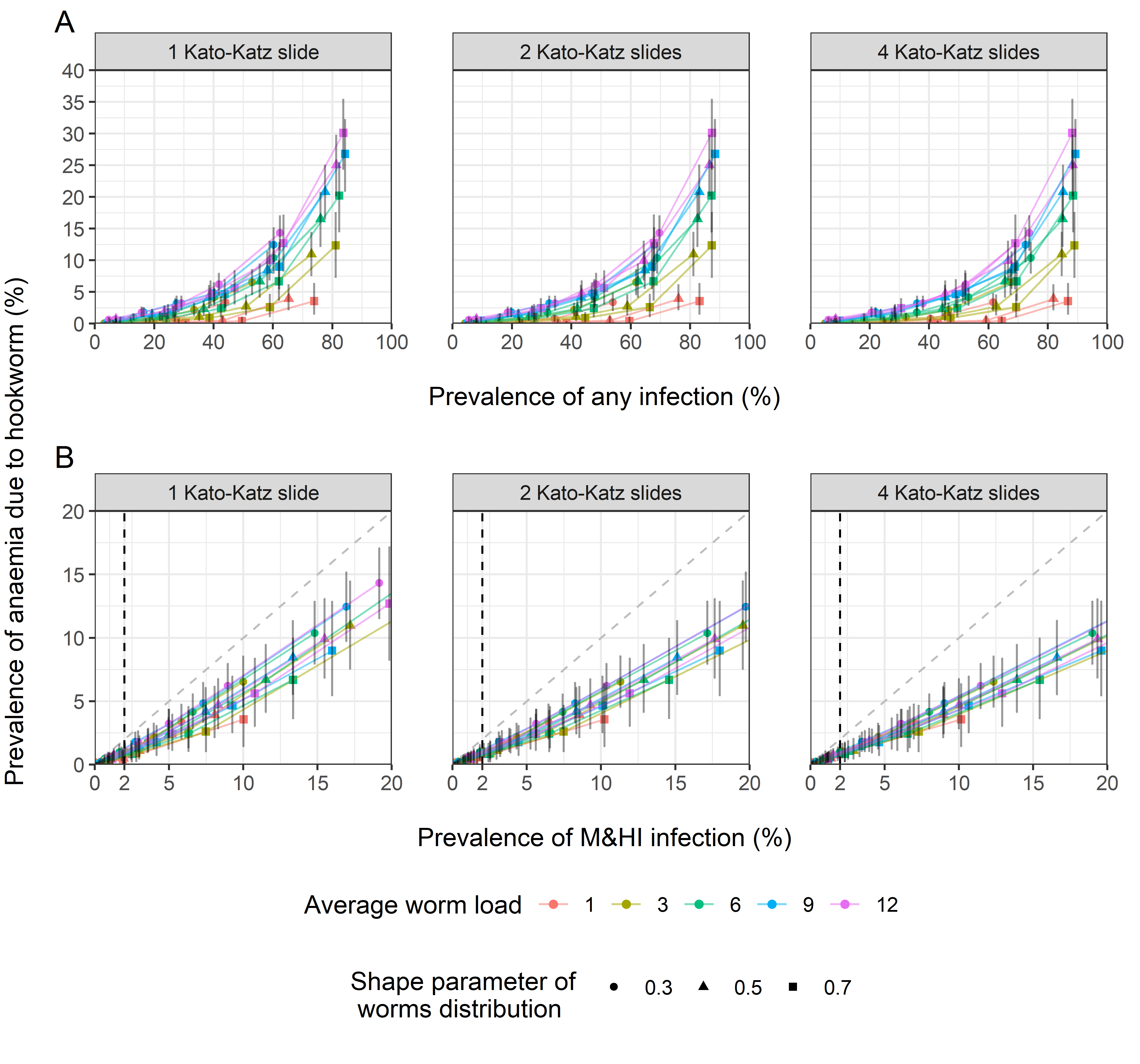
