## Supplementary material for "Appropriateness of the current parasitological control target for hookworm morbidity: a statistical analysis of individual-level data": S1 Table

| **Principle** | **What has been done to satisfy the principle?** | **Where in the manuscript is this described?** |
| --- | --- | --- |
| **Stakeholder engagement** | Work has been approved by the Bill and Melinda Gates Foundation as part of the NTD Modelling Consortium. The work has been presented at the 12^th^ European Congress on Tropical Medicine and International Health. | - |
| **Complete model documentation** | The statistical model implemented and used in this study is described in the manuscript. The full code for calibration of the model and simulations is available in a public repository.[1] | Methods section and References. |
| **Complete description of data used** | Data and parameters used are described in the manuscript. Data have been documented in previous publications.[2] The code for data selection and preparation is available in a public repository.[1] | Methods section and References. |
| **Communicating uncertainty** | We consider stochastic and parameters’ uncertainty in communicated results, via Bayesian Credible Intervals (BCIs) and Prediction Intervals (PIs). | All sections. |
| **Testable model outcomes** | The model outcomes can be tested by collecting individual data in different endemicity settings. | Discussion section. |
